# Sickle cell visualization *in vivo* in humans: microvascular occlusion formation and hemorheological indices

**DOI:** 10.1101/2025.09.27.25336804

**Authors:** Marisa M. Morakis, Luojie Huang, Gregory N. McKay, Sophie Lanzkron, Lydia H. Pecker, Nicholas J. Durr

## Abstract

Vaso-occlusion is a signature pathology of sickle cell disease (SCD). However, the lack of *in vivo* methods to observe individual blood cell dynamics in humans limits our understanding of occlusion formation mechanisms. We present a novel *in vivo*, non-invasive, label-free, and high-resolution imaging technique to study blood flow and sickled cell behavior in affected individuals. We used oblique back-illumination microscopy (OBM) to capture videos of 91.0 ± 42.3 sublingual capillaries in each of ten subjects with SCD before and after red cell transfusions and compared measurements to ten unaffected control subjects. With direct observation of blood cell activity, we identified microvascular occlusions initiated by red blood cells (RBCs) that adhered to the endothelium and caused mechanical vessel obstruction. Often, the RBCs were sickled. Then, in each observed vessel, we classified blood flow as fast, slow, or no flow, and counted adhered RBCs. Compared to controls, SCD subjects before transfusion had fewer fast-flowing vessels (48.7% vs. 77.7%, p=5.8×10^−4^), more no flow vessels (16.1% vs. 2.4%, p=0.0010), and more adhered RBCs (1.37 vs. 0.01 cells per vessel, p=0.0025). From before to after transfusion, SCD subjects’ microvasculature had increased fast-flowing (48.7% vs. 65.8%, p=0.0098) and decreased no flow vessels (16.1% vs. 6.0%, p=0.0039); adhered RBCs decreased (1.37 vs. 0.71 cells per vessel, p=0.043). These hemorheological indices captured transfusion-induced changes to vascular dynamics and events leading to microvascular dysfunction and occlusion in SCD. Our findings demonstrate the potential of OBM to study vaso-occlusion pathobiology, accelerate therapeutic evaluation, and personalize treatment strategies in people with SCD.

**Key Points:**

- Non-invasive *in vivo* blood imaging in humans reveals sickle cells initiate vaso-occlusion via endothelial interactions.
- *In vivo* hemorheological indices capture changes in blood flow related to sickle cell disease and transfusion response.

## Introduction

Sickle cell disease (SCD) is a monogenic autosomal recessive disorder caused by a mutation in the β-globin subunit of the hemoglobin protein. Sickle hemoglobin (HbS) polymerizes when deoxygenated, causing red blood cells (RBCs) to ‘sickle,’ becoming misshapen, stiff, brittle, and sticky.^1,2^ Vaso-occlusion, a hallmark SCD pathology, results from a combination of (1) the physical obstruction of capillaries by sickled RBCs, reticulocytes, white blood cells (WBCs), and platelets, and (2) chronic vasculopathy and inflammation.^3–7^ The mechanisms underlying vaso-occlusion are incompletely understood, including their initiation, duration, and dissipation. Besides the lack of comprehensive mechanistic insight, a challenge in developing therapies for SCD is that endpoints for clinical trials often rely on downstream clinical complications, including pain crisis frequency, which is subjective, or end-organ damage, which requires long-term monitoring. Biomarkers based on hematologic indices, such as hemoglobin or %HbS, are commonly used as clinical endpoints ^8,9^ but may not accurately indicate *in vivo* blood cell function.^10–12^ Identifying biomarkers that can consistently predict disease severity or specific complications for both clinical care and therapy trials has been challenging in SCD due to its highly heterogeneous phenotypes.

Most research on the pathobiology and hemorheology of SCD uses *in vitro* microfluidic models,^3,13–17^ *ex vivo* animal microvasculature,^18^ or transgenic mouse models that express human HbS.^5,19^ These methodologies allow visualization of flowing blood cells for rheological measurements such as viscosity, adhesion, and deformability of RBCs, which represent functional assessments of blood and are potential evaluation metrics of treatment efficacy. Rheological biomarkers reflect pathological differences between SCD genotypes,^20,21^ responsiveness to therapy,^22–24^ and risk for pain crises.^25^ Additionally, they help illuminate potential therapeutic targets.^16,26^ Visualization of flowing blood cells has also helped define our current understanding of vaso-occlusion mechanisms.^3,5^ However, knowledge of *in vivo* sickle cell behavior in humans has been limited by a lack of high-resolution, high-speed, label-free, and non-invasive imaging technologies. While widefield absorption-based imaging techniques have been applied to human SCD microcirculation, this approach lacks the contrast to distinguish individual RBCs from each other or discern characteristic sickle cell morphologies.^27–29^

We present the first study to directly observe individual blood cell dynamics and SCD vaso-occlusions *in vivo* in human microvasculature using oblique back-illumination microscopy (OBM). OBM is a phase contrast imaging technique for thick tissues ^30^ that allows the visualization of transparent features including high-resolution membrane contours and morphology of fast-flowing blood cells ^31,32^ as well as empty blood vessels. The oral mucosa is an optimal imaging site for non-invasive OBM imaging because it presents superficial capillaries without an absorbing melanin layer. We hypothesize that OBM in the ventral tongue will allow visualization of cellular events preceding vaso-occlusion, revealing the mechanism of occlusion formation. We also hypothesize that hemorheological analysis based on OBM imaging will capture hemodynamic changes in SCD compared to controls as well as in SCD subjects following red cell transfusion therapy.

## Methods

### Subjects and study design

We recruited ten adults with hemoglobin SS (N=9) or hemoglobin Sβ^0^ thalassemia (N=1) receiving chronic simple transfusions from the Johns Hopkins Sickle Cell Center for Adults. The protocol at the Center is to perform a draw-off for chronically transfused individuals presenting with a hemoglobin greater than 8 g/dL and then transfuse 2 units of packed RBCs, targeting a hemoglobin of 10 g/dL and ideally maintaining HbS fraction <30%. We recruited ten sex-matched adults with no known hematologic abnormalities as controls. Hemoglobin electrophoresis confirmed their hemoglobin AA genotypes.

For each subject with SCD, we collected ten 90-second OBM videos across two imaging sessions: five from the session immediately before transfusion and five from the session immediately after transfusion. For each control subject, we collected ten 90-second OBM videos in a single imaging session. Within two days before transfusion, subjects underwent a complete blood count, a hemoglobinopathy panel, and a reticulocyte count. We collected this same set of data immediately after imaging for post-transfusion sessions and control subjects. We compared subjects’ ages and hematologic indices with Student’s t-tests. Pre- to post-transfusion comparisons used paired tests, while SCD to control comparisons used unpaired tests. All subjects gave written informed consent, and data were collected under a Johns Hopkins University Institutional Review Board-approved protocol (IRB00264150) and in accordance with the Declaration of Helsinki.

### OBM imaging system and acquisition

We designed the OBM imaging system to acquire synchronized green and red channels. Green LED light (530 nm peak) provides high contrast of RBCs due to hemoglobin absorption. Red LED light (660 nm peak) has a longer scattering mean free path length in biological tissues than green light, which provides enhanced phase contrast. Each capillary video was acquired at 100 frames per second and 3.1 megapixels. A 20x 0.75NA multi-immersion objective lens (Nikon, CFI Plan Fluor 20XC MI) provided a 440 μm field of view and a theoretical resolution of 431 nm. To correct for the background intensity gradient across the field of view, we divided each image by a Gaussian-filtered version of itself (σ = 50 pixels). ^30,31^ To combine color channels, each frame is background corrected and registered, and the red channel is subtracted from green to preserve phase information from red and absorption information from green. We performed contrast adjustments to enhance visibility. We estimated velocity of individual cells in select cases by measuring the trajectory and the number of frames the cell took to travel this distance. Details of the optical system and hardware setup are published^31^ and reported in the Supplemental Methods and Supplemental Figure 1.

We modified a portable ophthalmic slit lamp microscope cart to accommodate the OBM microscope, retaining the existing forehead and chin rests for head stabilization, as well as the low-friction table for positioning the imaging system (Figure 1A). We 3D-printed a custom objective lens cap from stainless steel (Protolabs). The cap was surrounded by ports attached to a small vacuum pump to provide gentle suction to the ventral tongue for further stabilization (Figure 1B). During imaging, subjects placed their tongues on the objective cap (Figure 1C). Using a piezo objective scanner (Thorlabs PFM450E) for depth and the low-friction table for lateral translation, we positioned the microscope to focus on capillaries 0-200 μm deep into the ventral tongue surface. Once a shallow capillary bed was visible, subjects held still, and the vacuum was switched on when the video recording started.

**Figure 1.**
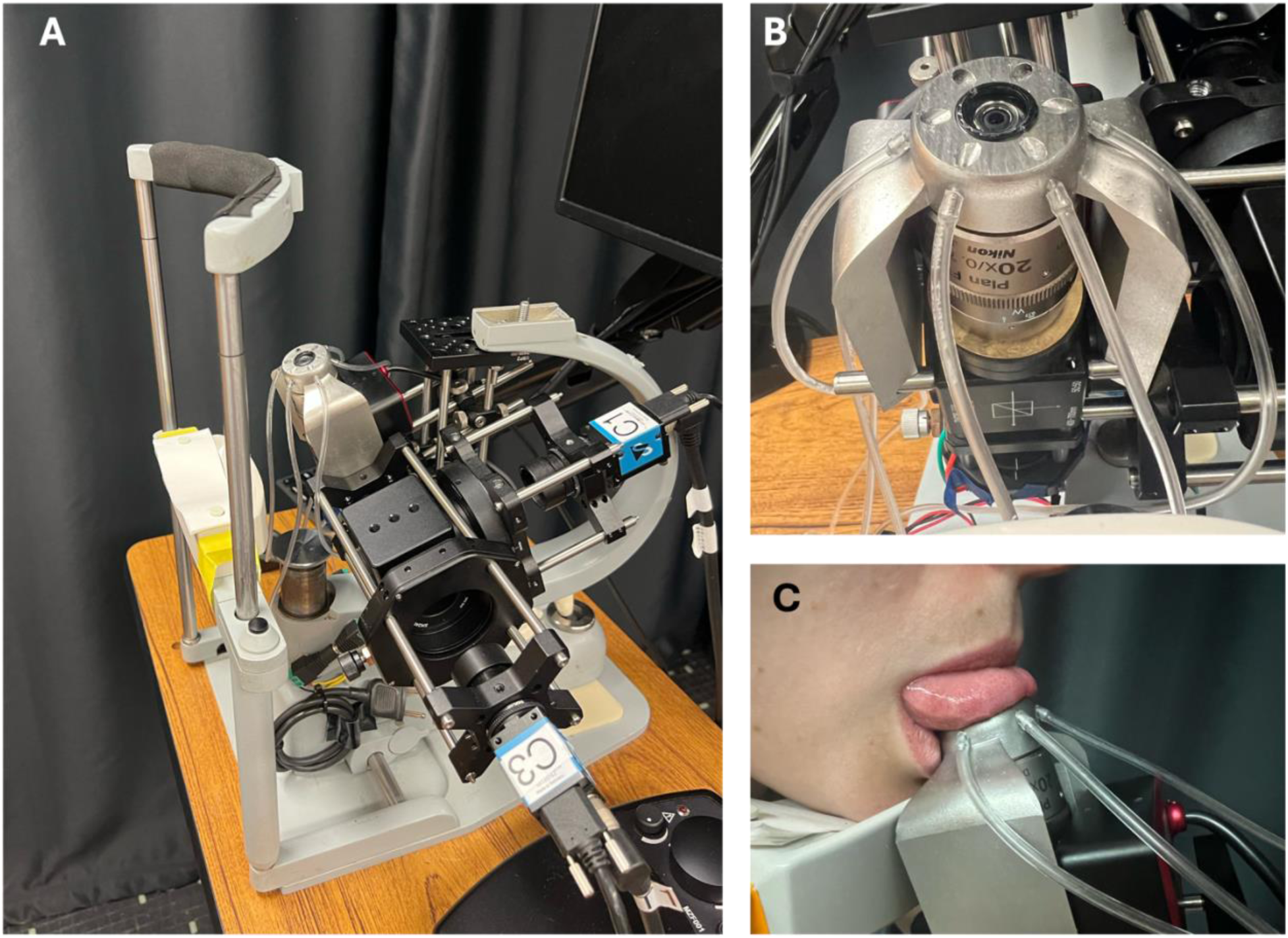
The oblique back-illumination microscope (OBM). (A) The repurposed slit lamp ophthalmoscope cart with attached OBM system includes forehead and chin rests to stabilize a subject’s head with respect to the imaging system and a low-friction mount for positioning the imaging system. (B) The custom objective lens cap applies gentle suction to the ventral tongue through six vacuum ports surrounding a glass imaging window. (C) An image of a volunteer researcher demonstrating the device use.

### Occlusion formation and dissipation characterization

We characterized microvascular occlusion formations observed in SCD subjects (Figure 2A; Videos 1 and 2). We defined an occlusion formation as a vessel transitioning from active RBC flow to a complete blockage by a visible buildup of cells. The initiation time point was the frame in which the first cell adhered at the occlusion site. The completion time point was the first frame in which the flow was completely stopped by cell buildup. The dissipation time point was the first frame in which RBCs began moving past the occlusion site. Occlusion formation time was the time from initiation to completion, and occlusion duration was the time from completion to dissipation.

**Figure 2.**
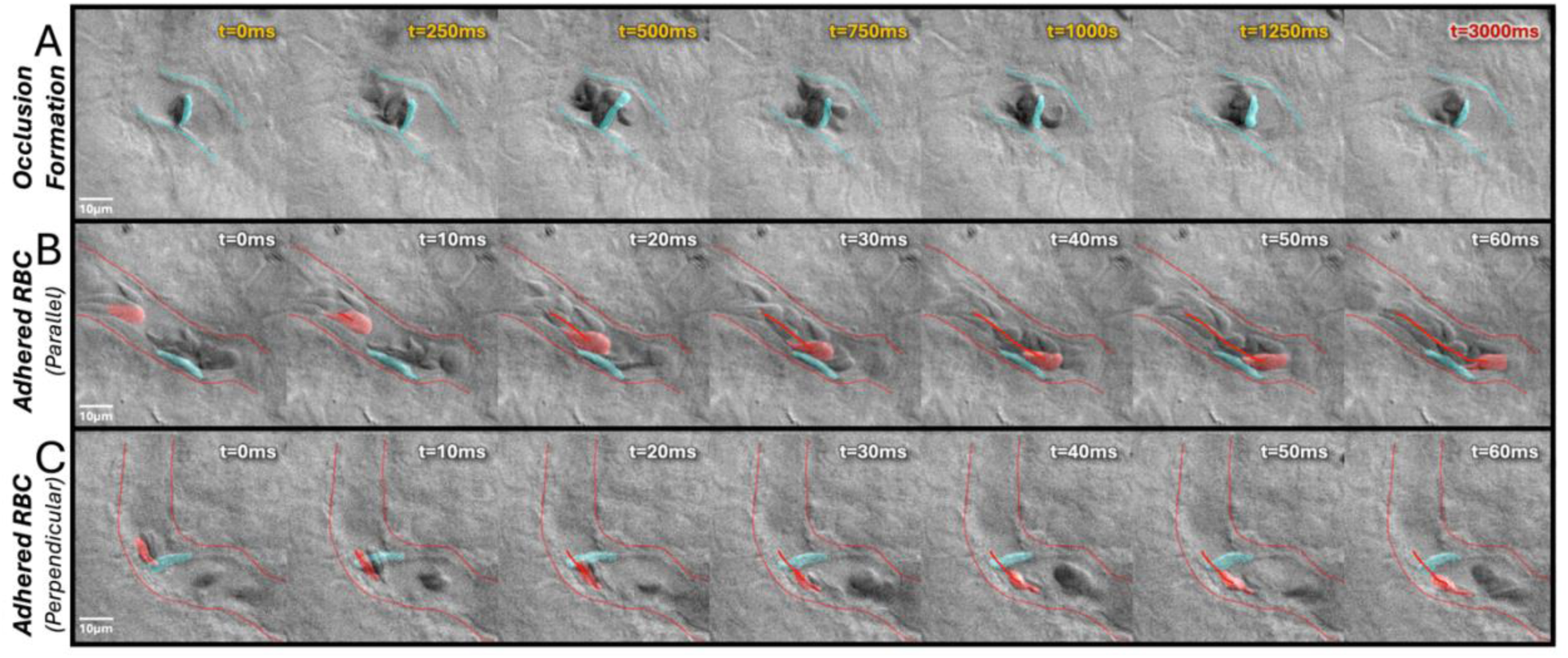
Occlusion formation and RBC adhesion examples. (A) Example of occlusion formation. An occlusion is initiated by a suspected sickle RBC (blue overlay) that adheres to the endothelium, followed by additional suspected sickle cells and RBCs adhering to the endothelium and/or the initial cell. Additional blood cells accumulate behind the adhered cells, blocking vessel flow. The occlusion happens over ∼1250ms, and no flow is seen for at least 1750ms. See Videos 1 and 2. RBCs (blue overlay) adhere to the endothelium along the direction of flow (B) or perpendicular to the direction of flow (C). Other RBCs (red overlay) flow past, with their trajectories marked by the red line. See Videos 3 and 4. Images are cropped combined red and green frames. Flow direction is left to right, and vessel walls are outlined. Time steps of 10 ms are written in white, 250 ms in yellow, and 1750 ms in red.

We further investigated the cells initiating observed occlusions. We defined occlusion-initiating cells as the first three cells to adhere in occlusion formation (if distinguishable). We evaluated occlusion-initiating cell morphology in comparison to control cells, defined as unaffected cells in no flow vessels of control subjects. Under OBM, we qualitatively identified “suspected sickled cells” based on elongated shape,^33^ higher phase contrast from increased hemoglobin fiber density, and lack of cell deformation between frames (for example, blue shaded cells in Figure 2). We also measured long axis length and aspect ratio, defined as the ratio of short to long axis length, of each occlusion-initiating cell to quantify size and shape. We performed 2D Gaussian regression to cluster the cells based on these two measurements. We further compared these measurements to those of control cells using a Student’s t-test.

### Hemorheological indices

Two graduate students with 3 years of experience in OBM and capillaroscopy (authors MMM, LH) performed rheological analysis manually. De-identification and random shuffling of videos before analysis ensured graders were blinded to subject group and condition. We categorized each vessel as either no flow, slow flow, or fast flow (Figure 3; Videos 5 and 6). We defined *no flow* as no visible movement of blood cells in the vessel throughout the video, which included both cell buildups and empty vessels; *slow flow* as partially occluded, intermittent, sluggish, or sparse blood flow; and *fast flow* as continuous, smooth flow with abundant blood cells. We presented flow categorization results per session as the percentage of vessels in each category out of all observed vessels, where each session consisted of 5-10 90-second videos. For robust classification, only vessels visible for at least 50 frames (0.5 seconds) were included. Additionally, we quantified RBC adherence to the endothelium, defining adhered RBCs as RBCs that remained immobile within the vessel for at least 20 frames (0.2 seconds) while other cells flowed past (Figure 2B,C; Videos 3 and 4). We normalized adhered RBC counts to the total number of observed vessels per session.

**Figure 3.**
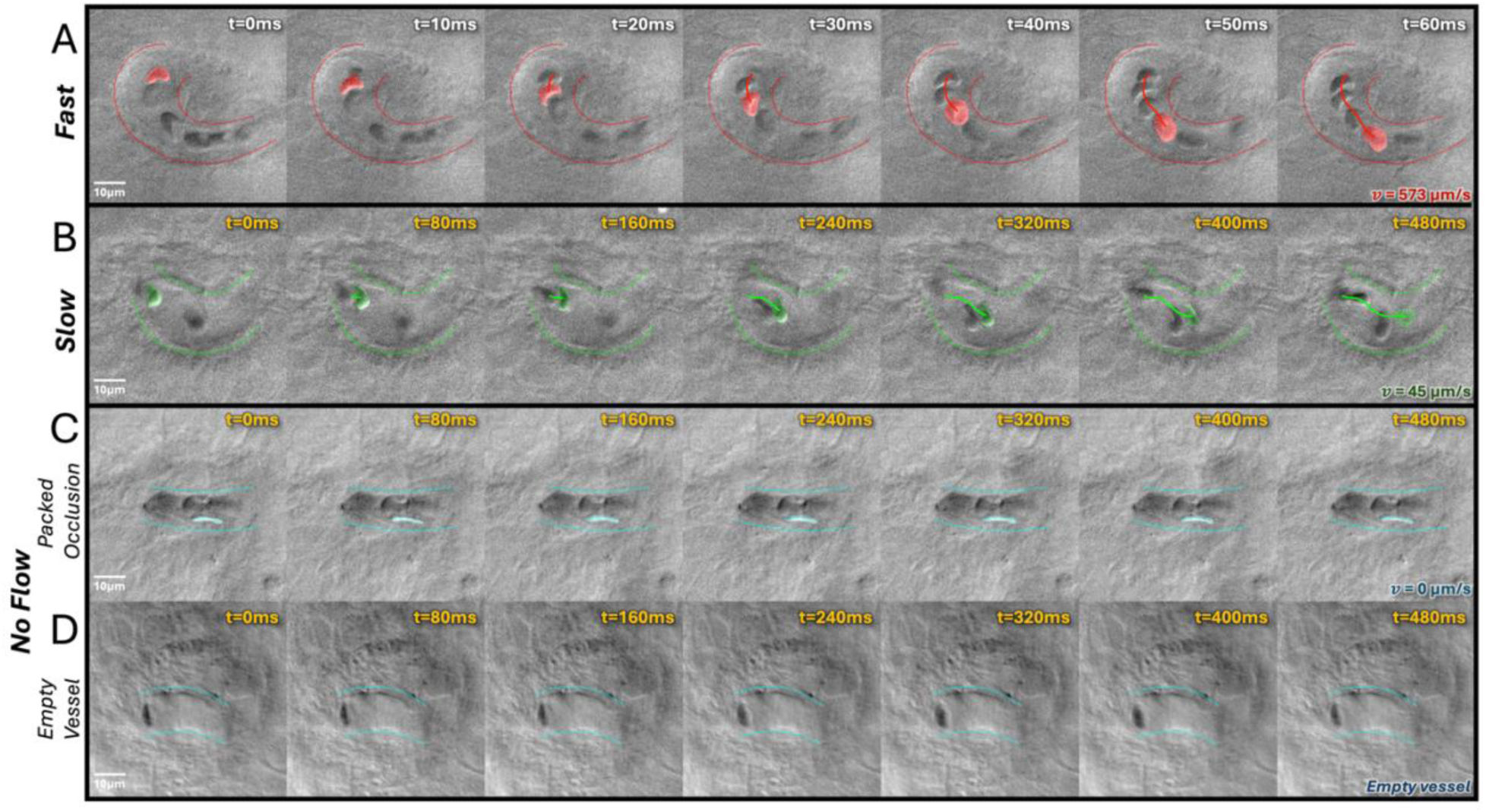
Comparison of different flow categories. (A) ***Fast flow*** with a blood cell flowing at 574 μm/s. See Video 1. (B) ***Slow flow*** with a blood cell flowing at 45 μm/s. The cell traveled a similar distance to the cell in (A), but over a significantly longer period of time. See Video 2. (C) ***No flow*** as a packed occlusion. An RBC involved in an occlusion (blue overlay mask) remains stationary throughout the observation period. (D) ***No flow*** as an empty vessel. An empty capillary is occluded either upstream or downstream from the visible segment, with one stationary RBC visible. Images are cropped combined red and green frames. Flow direction is left to right, and vessel walls are outlined. Shaded overlays on blood cells indicate the same cell tracked across adjacent frames, and the line indicates its trajectory. Time steps of 10 ms are written in white and 80 ms in yellow.

To assess inter-rater reliability (IRR), we compared the evaluations of both graders on the same set of 191 vessels from a randomly selected subject, which included both pre- and post-transfusion sessions. We calculated the IRR with a weighted Cohen’s kappa for ordinal variables with quadratic weighting, where a kappa value of at least 0.6 represents substantial agreement and at least 0.8 represents near-perfect agreement.^34^ For appropriate statistical tests of compositional data, we applied a centered log-ratio (CLR) transform to flow category percentages, x: CLR(x_i_) = log(x_i_/g(X)), g(X) = ^3^Ö(x_1_*x_2_*x_3_).^35^ We used non-parametric Wilcoxon signed rank tests to compare paired pre- and post-transfusion sessions and Mann-Whitney rank sum tests to compare SCD and control subjects. We defined statistical significance as α=0.05 for all tests.

To assess relationships between hemorheological indices and standard SCD laboratory biomarkers—%HbS, absolute reticulocyte count (ARC), and hemoglobin—we used simple linear regression. We calculated goodness of fit with the coefficient of determination (r^2^), as well as 95% confidence intervals for the slope and intercept of each regression line.

## Results

### Subject characteristics

There was no significant difference between groups in subject age (SCD 34.50 ± 14.59 years vs. controls 26.70 ± 4.06 years, p=0.40). Recruitment of SCD subjects was not sex-selective, but only females participated. No subjects experienced pain crises or other adverse events during or after participation in the study. Table 1 presents hematologic indices in control subjects and SCD subjects pre- and post-transfusion. Mean hemoglobin levels increased after transfusion in SCD subjects (8.64 ± 1.35 g/dL to 10.11 ± 1.29 g/dL, p=5.8×10^−5^). Mean hemoglobin in control subjects was higher than post-transfusion SCD levels (13.33 ± 0.79 g/dL, p = 2.6×10^−6^). Mean platelet counts decreased after transfusion in SCD subjects (451.50 ± 212.45 K/μL to 393.10 ± 173.62 K/μL, p=0.0028). In the four subjects where post-transfusion HbS% was measured, HbS% decreased after transfusion (45.18 ± 18.37 to 32.53 ± 13.04, p=0.018).

**Table 1.** Hematologic indices of subjects.

| Hematologic index | SCD<br>N = 10<br>Mean ± SD | SCD<br>N = 10<br>Mean ± SD | Control<br>n = 10 | P value | | |
| --- | --- | --- | --- | --- | --- | --- |
|  | Pre-Transfusion | Post-Transfusion |  | Pre-Post | Pre-Control | Post-Control |
| **Hb, g/dL** | 8.64 ± 1.35 | 10.11 ± 1.29 | 13.33 ± 0.79 | 5.8x10<sup>-5***</sup> | 2.1x10<sup>-8***</sup> | 2.6x10<sup>-6***</sup> |
| **Hct, %** | 25.46 ± 3.92 | 29.53 ± 3.93 | 39.57 ± 2.04 | 1.4x10<sup>-4***</sup> | 7.6x10<sup>-9***</sup> | 1.1x10<sup>-6***</sup> |
| **RBC, 10<sup>6</sup>/μL** | 2.83 ± 0.46 | 3.36 ± 0.46 | 4.34 ± 0.27 | 2.2x10<sup>-5***</sup> | 4.8x10<sup>-8***</sup> | 1.6x10<sup>-5***</sup> |
| **MCV, fL** | 90.09 ± 2.85 | 87.91 ± 2.53 | 91.24 ± 3.17 | 0.0012** | 0.40 | 0.018* |
| **RDW, fL** | 21.29 ± 2.78 | 19.42 ± 2.16 | 12.37 ± 0.41 | 3.8x10<sup>-5***</sup> | 8.4x10<sup>-9***</sup> | 7.4x10<sup>-9***</sup> |
| **WBC, K/μL** | 12.06 ± 3.22 | 11.11 ± 3.59 | 6.50 ± 0.82 | 0.41 | 4.9x10<sup>-5***</sup> | 9.3x10<sup>-4***</sup> |
| **Neutrophils, K/μL** | 7.27 ± 2.64 | 5.88 ± 1.93 | 3.73 ± 0.74 | 0.23 | 6.8x10<sup>-4***</sup> | 0.0040** |
| **Platelets, K/μL** | 451.50 ± 212.45 | 393.10 ± 173.62 | 260.50 ± 31.91 | 0.0028** | 0.012* | 0.029* |
| **ARC, K/μL** | 413.10 ± 173.99<br>(N=9) | 373.98 ± 72.53<br>(N=4) | 59.39 ± 16.36 | 0.51 (N=3) | 6.4x10<sup>-6***</sup> | 1.1x10<sup>-8***</sup> |
| **HbS, %** | 39.52 ± 18.09 | 32.53 ± 13.04<br>(N=4) | NA | 0.018* | NA | NA |
| **HbF, %** | 3.10 ± 2.24 | 3.68 ± 2.10<br>(N=4) | NA | 0.037* | NA | NA |

Hb indicates hemoglobin; Hct, hematocrit; MCV, mean corpuscular volume; RDW, red cell distribution width; HbF, fetal hemoglobin; and NA, not applicable. \* P<0.05; \*\* P<0.01; \*\*\* P<0.001

### Occlusion formation and dissipation

We observed 48 total occlusion formations (N=37 pre-transfusion, N=11 post-transfusion). While all 10 SCD subjects exhibited occlusions based on the existence of no flow vessels, new occlusions were observed forming during the observation period in 8 out of 10 subjects. Capillary diameters of vessels where these occlusion formations occurred were 11.7 ± 1.93 μm.

All 48 observed occlusion events were initiated by adhesion of RBCs to the endothelium. Example occlusion formations are shown in Video 1 (with representative frames in Figure 2A) and in Video 2. Subsequent RBCs adhered to the initial adherent cell and adjacent endothelial cells. Occlusion-initiating cells had more elongated shapes than control cells (aspect ratio 0.31 ± 0.14 vs. 0.61 ± 0.20, p=1.3×10^−17^). Distributions of occlusion-initiating cell aspect ratios and long axis lengths were largely distinct from those of control cells (Figure 4B). Suspected sickle cells based on qualitative classification were the longest and had the lowest aspect ratios, showing very little overlap with control cells: suspected sickle cell aspect ratios were mostly below 0.3, while control cells were mostly above this threshold. Forty of the 48 occlusion formations involved at least one suspected sickle cell in the three occlusion-initiating cells.

**Figure 4.**
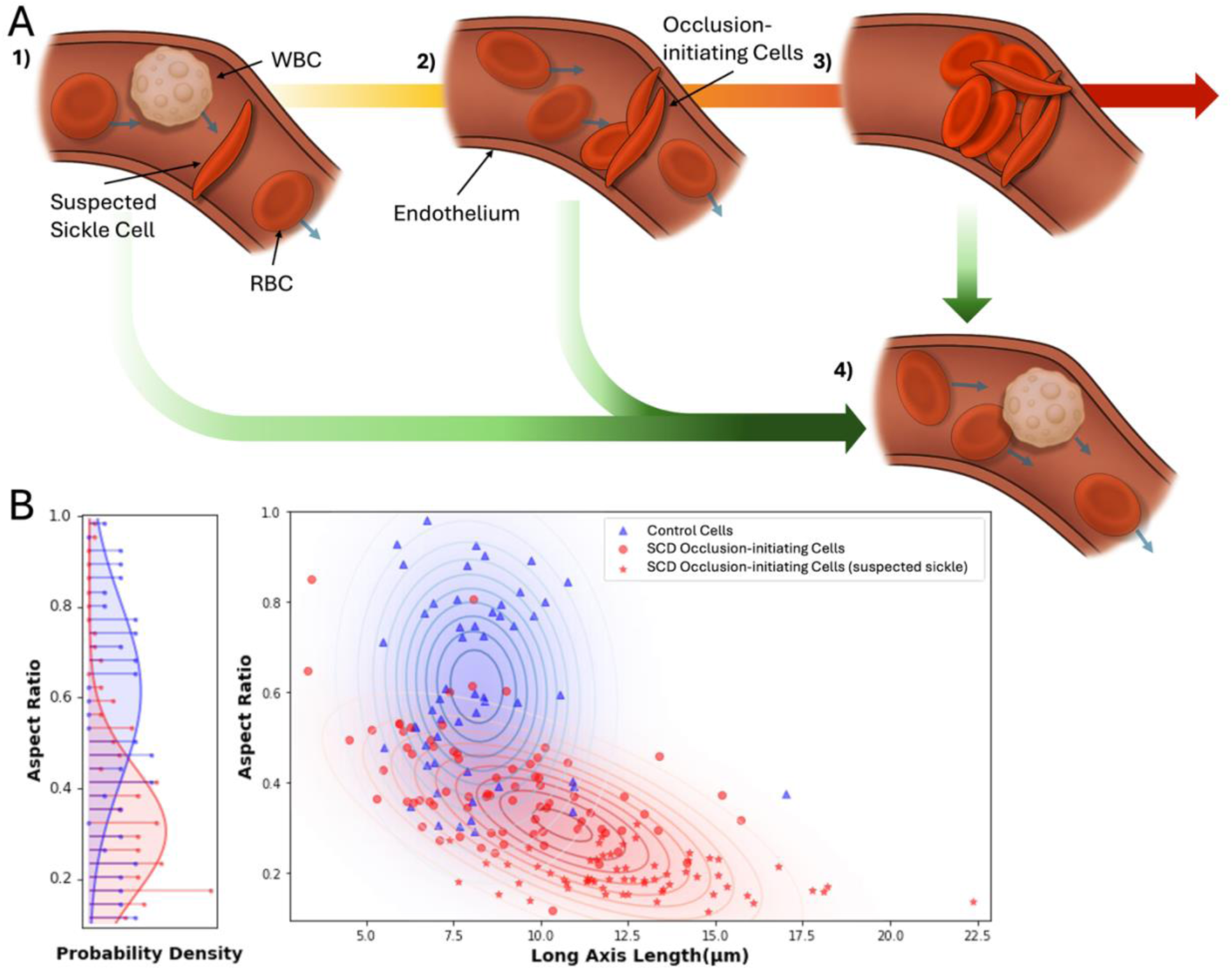
Observed mechanism for SCD vaso-occlusion at the capillary level. (A) Vaso-occlusion was initiated by a sickle RBC adhering to the endothelium (1). Although red cell adhesions were often isolated and would eventually detach (1 to 4), they also initiated the buildup of cells. As other cells flowed past, other occlusion-initiating RBCs adhered to the initial cell or the surrounding endothelium (2). These buildups of multiple cells were sometimes seen dissipating (2 to 4) before an occlusion formed. Finally, buildups became large enough to trap all other cell types, resulting in a full vaso-occlusion with no flow in the vessel (3). Occlusions were sometimes seen dissipating (4). (B) The left pane is the probability density functions of aspect ratio for control and occlusion-initiating cells, overlaid with a Gaussian fitting. The right panel shows the 2D distributions of control and occlusion-initiating cells along aspect ratios and cell long-axis lengths, with suspected sickle cells marked by stars. A 2-dimensional Gaussian fitting is shown for each group.

Following the occlusion-initiating cells, more RBCs and occasional WBCs accumulated until the capillary became occluded. The observed mechanism is depicted in Figure 4A. The time course of the formation and dissipation processes varied widely. For the 20 cases where full occlusion formation time was measurable, occlusions took between 0.81 and 52.32 seconds to form. For 11 cases where the occlusion dissipation was also observed, the occlusion duration ranged from 0.45 to 60.05 seconds. For all other cases, occlusions moved out of the field of view during observation and duration was not measurable.

### Hemorheological indices

For a comprehensive study of hemorheological parameters related to the vaso-occlusion mechanisms we observed, we counted adhered RBCs and quantified vessels in each of three flow speed categories: no flow vessels represented overall occlusion frequency, slow flow vessels potentially indicated partial or developing occlusions, and fast flow vessels represented unobstructed vessels. Figure 5 shows flow speed percentage distributions in each experimental group. A mean of 91.0 ± 42.3 vessels were observed per session. Compared to controls, pre-transfusion SCD subjects had lower percentages of fast flow (48.7 ± 17.39% vs. 77.8 ± 11.54%, p=5.8×10^−4^) and higher percentages of no flow (16.1 ± 10.58% vs. 2.4 ± 2.01%, p=0.0010) vessels. From pre- to post-transfusion, SCD subjects’ percentages of fast flow vessels increased (48.7 ± 17.39% to 65.8 ± 17.39%, p=0.0098) and percentages of no flow vessels decreased (16.1± 10.58% to 6.0 ± 5.40%, p=0.0039). There were no differences in flow speed distributions between post-transfusion SCD subjects and controls. IRR testing yielded a weighted Cohen’s kappa of 0.825, indicating strong agreement of flow categorization between two graders. Additional IRR results can be found in Supplemental Figure 2.

**Figure 5.**
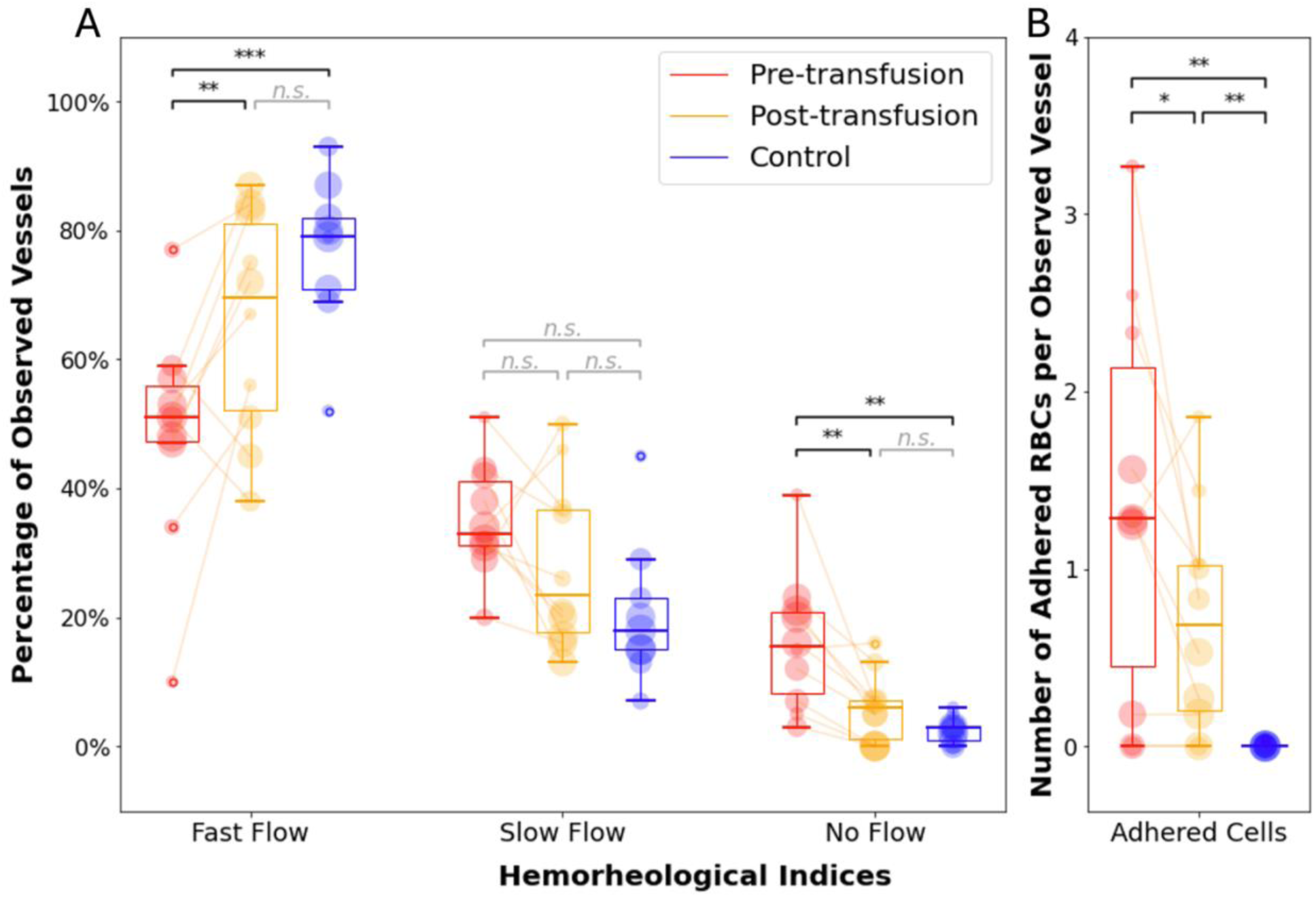
In-vivo hemorheological indices between pre- and post-transfusion SCD subjects, and controls. Boxplots display the distribution of hemorheological indices in each group. (A) Percentages of observed vessels in each imaging session assigned to each flow category: fast flow, slow flow, or no flow. (B) Number of adhered RBCs per observed vessel in each imaging session. Boxes represent the interquartile range, the centerline marks the median, and whiskers extend to the farthest data point within 1.5x the interquartile range from the box. Data points beyond this range are marked by circles. Each data point represents a measurement from a single imaging session, and its size indicates a kernel density estimate of the data distribution. Lines between data points show intra-subject changes pre- to post-transfusion. * P<0.05; ** P<0.01; *** P<0.001; n.s. not significant.

In SCD subjects, we observed 1.37 ± 1.11 adhered RBCs per vessel pre-transfusion, which decreased to 0.71 ± 0.63 adhered RBCs per vessel post-transfusion (p=0.043). Adhered RBCs were rare in controls; we observed 7 adhesion events in 1,047 total vessels.

There were moderate correlations between OBM-derived hemorheological indices and hematologic indices (Figure 6). Pre-transfusion %HbS was positively correlated with no flow (r^2^=0.50) and negatively correlated with fast flow (r^2^=0.43). ARC had little correlation with flow metrics but had a weak positive correlation with adhered RBCs pre-transfusion (r^2^=0.26). Hemoglobin showed a slight negative correlation with no flow, both pre- and post-transfusion (r^2^=0.17, pre-transfusion; r^2^=0.19, post-transfusion), and with adhered RBCs post-transfusion (r^2^=0.36).

**Figure 6.**
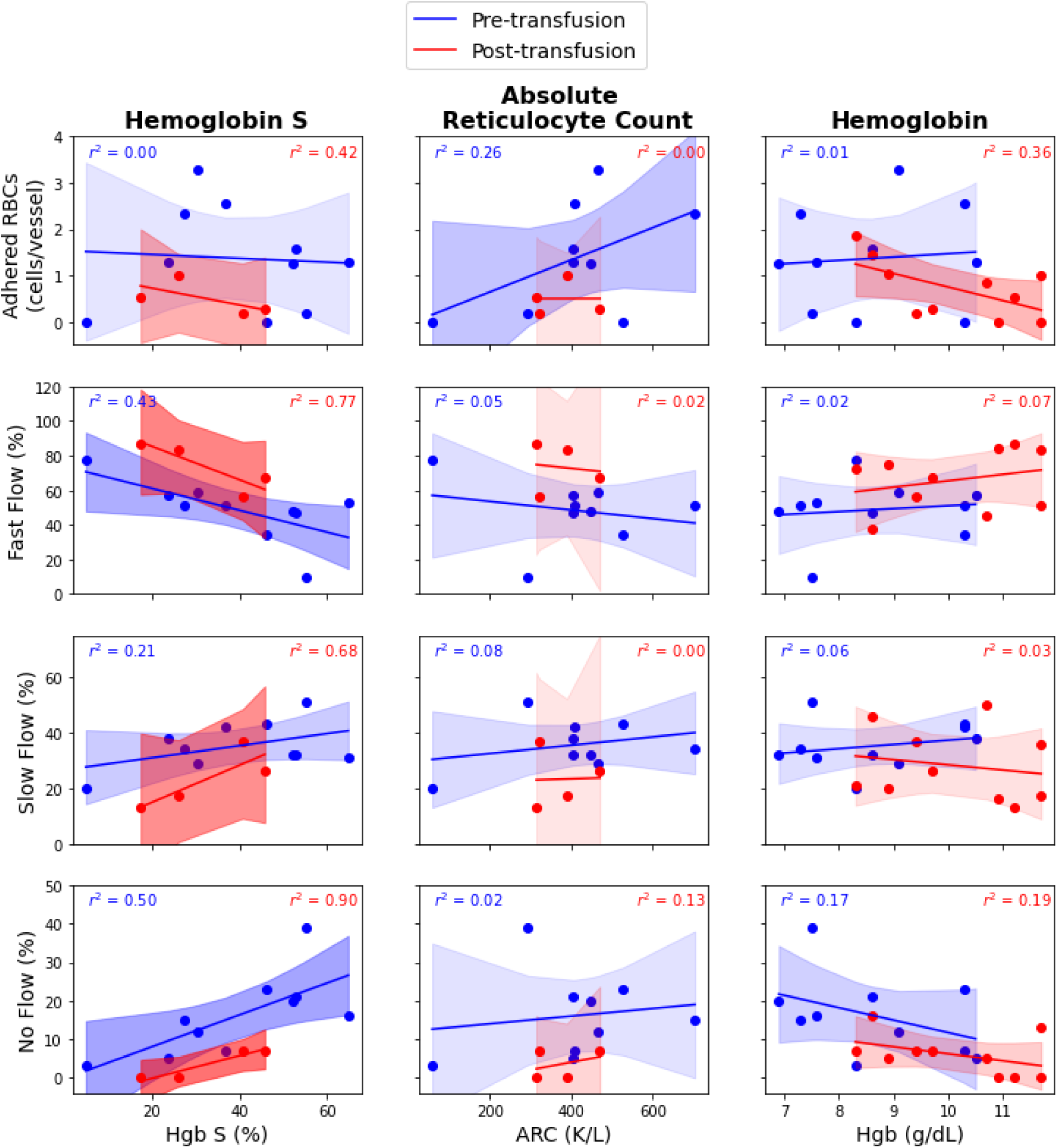
Correlations between hematologic indices and hemorheological indices. Pre-transfusion data points and linear regression are in red, while post-transfusion data is in blue. r^2^ values and 95% confidence intervals for slope and intercept are displayed. The confidence interval opacity corresponds to the r^2^ value.

## Discussion

To the best of our knowledge, this study is the first to visualize individual blood cell behavior, including vaso-occlusion formation, in the microvasculature of humans with SCD with cellular resolution. Phase contrast from OBM allows visualization of cell membrane contours, supporting distinction of cells when clustered and morphologic characterization of individual sickled RBCs. Direct observation with OBM shows that RBCs, which were often suspected sickle cells with elongated shapes, solely initiate occlusion through adherence to the microvasculature endothelium with subsequent integration of additional RBCs and WBCs. Additionally, with proposed *in vivo* hemorheological indices, we observed significant differences in blood flow behavior and adhered RBC counts in subjects with SCD compared to controls and after transfusion. These data offer insight into pathobiological mechanisms of SCD and suggest that OBM may be a useful tool in appraising response to existing and novel therapeutic strategies.

This study offers a new perspective on the mechanism of SCD vaso-occlusion formation. While RBC involvement in vaso-occlusion is uncontested, the precise order of events leading up to vaso-occlusion is unclear.^36–38^ Based on *in vitro*^1,39^ and *ex vivo*^18^ experiments, an early theory on occlusion formation is that adhesion of deformable RBCs to the endothelium is followed by trapping of dense sickle RBCs that occlude vessels.^40^ In contrast, intravital microscopy in murine SCD models later observed that sickle cell interactions with adhered leukocytes cause occlusions in vessels 20-25 μm in diameter,^5^ suggesting the inciting event in vaso-occlusion is WBC-endothelial adhesion. Encompassing both models, one idea proposes that occlusions in smaller vessels or during steady state are RBC-initiated, while occlusions in larger vessels or those triggered by inflammation are WBC-initiated.^37,41^ Since our *in vivo* observations were in smaller capillaries only (11.7 μm in diameter) and subjects were not experiencing pain crises, our findings may be consistent with this proposal. Furthermore, we observed some occlusions forming or dissipating in seconds and lasting up to a minute, though the duration of all observed occlusions could not be measured. The association of observed processes, especially these transient occlusions,^42^ with pain or ischemic damage remains unknown. Knowledge of the cellular processes driving formation of occlusions related to vaso-occlusive events, like pain or acute chest syndrome, may inform the molecular targets of potential disease-modifying therapies and guide personalized strategies. The mechanisms described here suggest that interventions targeting RBC sickling and adhesive behavior^43,44^ may be effective in these individuals, since adhered sickled RBCs appeared to be inciting factors in vaso-occlusion.

OBM-derived hemorheological indices demonstrate relevance to known hematologic features of SCD, mirroring validated microfluidic measurements of an individual’s propensity for adhesion and occlusion. *In vivo*, high HbS% (>40%) was associated with high no flow frequency, suggesting that the prevalence of sickle hemoglobin leads to more frequent occlusions. Man et al ^45^ proposed the occlusion index to measure the percentage of a capillary network on a microfluidic chip that was occluded by a SCD blood sample. Like no flow frequency, occlusion index increased in SCD compared to controls and correlated with HbS%.^23,45^ Adhered RBCs increased with ARC in our study. Sickle reticulocytes have enhanced adhesive properties; possibly, some of the adhered RBCs we observed were reticulocytes. Many other *in vitro* assays are designed to study adhesion in SCD and have also found associations between adhered RBCs and ARC.^21,46^ As evidenced by these parallels, OBM may be well-suited and advantageous for use in applications where these microfluidic assays for SCD hemorheology are already utilized, including evaluating response to treatments and investigating the basis of genetic and phenotypic variability.^16,20–22,24,26,46,47^

Hemorheological indices may be even more informative of microvascular health in SCD than standard hematologic analysis. There is likely more complexity in microvascular function and cellular interaction that hematologic indices, including protein biomarkers, cannot fully capture.^12,48^ In this study, correlations between hemorheological and hematological indices were all relatively weak, but improved hemorheology after transfusion and in controls compared to SCD subjects was still captured clearly.

Previous studies of SCD microcirculation show partial agreement with our hemorheological findings, though with key differences attributable to methodological and subject differences from our chronically transfused adult population. These differences underscore the improved sensitivity of our study in evaluating microvascular flow due to the design of quantification metrics and the phase contrast of OBM. Widefield absorption-based imaging and either average flow velocity or microvascular flow index (MFI), a metric based on subjective labeling of flow speed per image quadrant and averaging the result,^49–51^ have been used to quantify microcirculatory flow speed. However, averaging velocity across vessels may conceal the underlying redistribution of flow speeds we observed, and MFI may be dominated by more common flow categories to conceal differences in less frequent no flow vessels (Figure 5A). Prior comparisons of flow speeds in controls and SCD subjects disagree with each other. The conjunctival microvasculature demonstrated higher average blood flow velocity in controls compared to SCD subjects with severe symptoms, ^27^ consistent with our findings of increased fast flow vessels and decreased no flow vessels in SCD subjects, though no significant changes were reported in SCD subjects with nonsevere symptoms in that study. Other studies of sublingual microvasculature reported no changes in flow speeds between controls and SCD subjects in steady state or crisis.^28,29^ However, in a closer examination of flow categories that make up MFI, the number of vessels in the no flow and intermittent flow categories correlated with hemoglobin levels.^28^ This is evidence for the value of exploring the distributions of flow speed categories rather than aggregated measures.

Phase contrast provided by OBM also offers advantages over traditional absorption-based imaging used in previous SCD microcirculation studies. Absorption-based imaging captures lower-resolution signal from deeper tissue structures, whereas OBM only captures higher-resolution, optically sectioned images of the most superficial capillaries less than 200 μm deep. High resolution and phase contrast in OBM reveal individual cell boundaries even when cells are closely clustered, facilitating a more accurate perception of flow speed. OBM also enables the identification of empty capillaries (Figure 3D) and thus improves the estimation of no flow vessels. In another study, pediatric SCD patients’ conjunctival microvasculature contained more vessels after exchange transfusions than before.^52^ Increased vessel density likely indicates renewed perfusion of previously occluded vessels that were invisible under absorption-based microscopy but can be directly detected with OBM. These results agree with ours, since we also observed a decrease in no flow vessels after transfusion. However, in the same study, averaged flow velocities decreased slightly after transfusions, in contrast with the increase in fast flowing vessels we observed. Like with the studies described above, this discrepancy can be attributed to the differing methodology of velocity averaging.

Despite the advantages of our hemorheological indices, the flow patterns in the “slow” category remained difficult to interpret across groups. Slow flow showed little change between controls and SCD subjects pre- and post-transfusion, possibly due to viscosity increases after transfusion that could have confounded the improvements in flow speed from improved oxygenation of healthy RBCs.^53^ This was also potentially due to the broad definition used for subjective classification of slow vessels, encompassing slow speed, intermittent flow, and partial occlusion. Leveraging automatic quantification and further distinction in the “slow” category could help to disentangle compound impacts of sickle cell adhesion, occlusion, and increased blood viscosity induced by transfusion.

Limitations of this study should be acknowledged. The sample was small and all female. The historical exclusion of women in medical research has created detrimental knowledge gaps,^54^ so it is important to avoid repeating such imbalances. Though imaging in the ventral tongue maximized OBM contrast in superficial capillaries, stability and generalizability of the imaging site can be improved. Residual background motion prevented continuous imaging of the same vessels during our limited recording time of 90 seconds per video. As a result, occlusion formation and dissipation dynamics happening on a longer time scale were not observable. It is also unknown whether the imaging procedure—placing the tongue on a metal objective cap outside the mouth and applying gentle suction—impacts the microvascular behavior. Finally, the correlation between the superficial microvasculature and the microcirculation in other parts of the body has yet to be established, which is critical for exploring the influence of these findings on end organs such as the brain, spleen, or kidneys.

Future work will expand the study population and data collection to verify the mechanisms and measurements described here in a broader and more diverse population, including more males. Pain crisis and organ damage outcomes will be collected in this larger cohort to help clinically validate OBM-derived hemorheological indices as biomarkers for disease complications. Other oral mucosa sites, such as the inner lip,^32^ will be explored for collecting longer and more stable videos. Stable videos would enable future automated analysis,^55^ including capillary segmentation, cell tracking, and cell classification for a more thorough sickle cell morphologic analysis and flow quantification. Morphologic analysis of sickle cells can be further improved by incorporating volumetric imaging techniques such as high-speed scanning or multi-focus microscopy,^56^ since 3-dimensional shapes could not be unambiguously characterized under 2-dimensional OBM.

Here, we demonstrate the potential of OBM as a non-invasive and label-free tool in monitoring SCD. We show that sickled RBCs initiate vaso-occlusions in previously unseen SCD dynamics. We measure *in vivo* hemorheological properties in SCD microvasculature before and after transfusions, demonstrating an improved hemorheological state with faster flow, fewer occlusions, and fewer adhered RBCs after transfusion therapy. This approach offers a novel *in vivo* view of blood cell behavior and an opportunity to elucidate aspects of SCD pathobiology that are incompletely understood. With a better understanding of the clinical outcomes associated with the indices and processes observed here, OBM has the potential to guide future drug development, personalized therapy management, and evaluation of treatment efficacy.

## Data Availability

For original data, please contact.

## Acknowledgements

This work was supported by research funding from the Bill & Melinda Gates Foundation (INV-006005), NIH Tech Accelerator Challenge, and Fifth Generation to N.J.D.; from the NSF Graduate Research Fellowship Program to M.M.M.; and from NIH/NHLBI K23HL146841, NIH/NHLBI U01 HL156620-01, the Food and Drug Administration’s Office of Women’s Health and Center of Excellence in Regulatory Science and Innovation, the Mellon Foundation, Affimmune, and Novartis to L.H.P.

The authors thank the patients at the Johns Hopkins Sickle Center for Adults and volunteers for their participation in this study. We also acknowledge the invaluable contributions of our research assistants, especially Lauren Anthony, Rhonda Miller, Kora Coker, and Jayla Scott.

## Authorship

### Contribution

N.J.D., L.H.P., M.M.M., and L.H. designed the study; M.M.M., L.H., G.N.M., and N.J.D. built the microscope; M.M.M. and L.H. collected and analyzed the data; M.M.M wrote the manuscript; all authors reviewed and edited the manuscript; and N.J.D and L.H.P. oversaw the project.

