## Supplemental Data for "Sickle cell visualization *in vivo* in humans: microvascular occlusion formation and hemorheological indices"

### Methods

The OBM optical system is similar to the one described by McKay et al.[1] It uses LEDs imaged onto the focal plane as light sources offset from the detection axis rather than an external optical fiber placed adjacent to the objective lens, as in traditional OBM.[2] In this implementation, the LED board (Luxeon Star LEDs, Saber Z4 20 mm Square Color Mixing Array) contains two LEDs with wavelengths of 530 nm (30 nm spectral half width) and 655 nm (20 nm spectral half width) to acquire synchronized absorption-weighted and phase-weighted videos. The LEDs are imaged through an aspheric collimating lens (Thorlabs, ACL1512U-A) and the objective lens (Nikon, CFI Plan Fluor 20x/0.75 MImm) onto the objective's focal plane. In the object space, the LED images are 353 μm apart and oriented on opposite sides of the imaging FOV, providing two opposing directions of phase gradient contrast. Back-scattered light is collected by the objective and reflected by a 50:50 non-polarizing beamsplitter (Thorlabs, CCM1-BS013) to a tube lens (Thorlabs, AC254-200-A-ML). A dichroic mirror then splits the image formed by the tube lens into red and green color channels, which are recorded synchronously with two CMOS cameras at 100 frames per second (The Imaging Source, DMK 33UX252) triggered by an Arduino.

The objective lens cap (Figure 1) is fixed with respect to the imaging system and holds a no. 2 coverslip. Six vacuum ports surround the coverslip slot and connect to a compact vacuum pump (Pacum, Masterspace) which applies gentle suction. A piezo objective scanner is used to scan the objective lens along the optical axis over a range of 450 μm, allowing focusing on different planes inside the capillary bed.

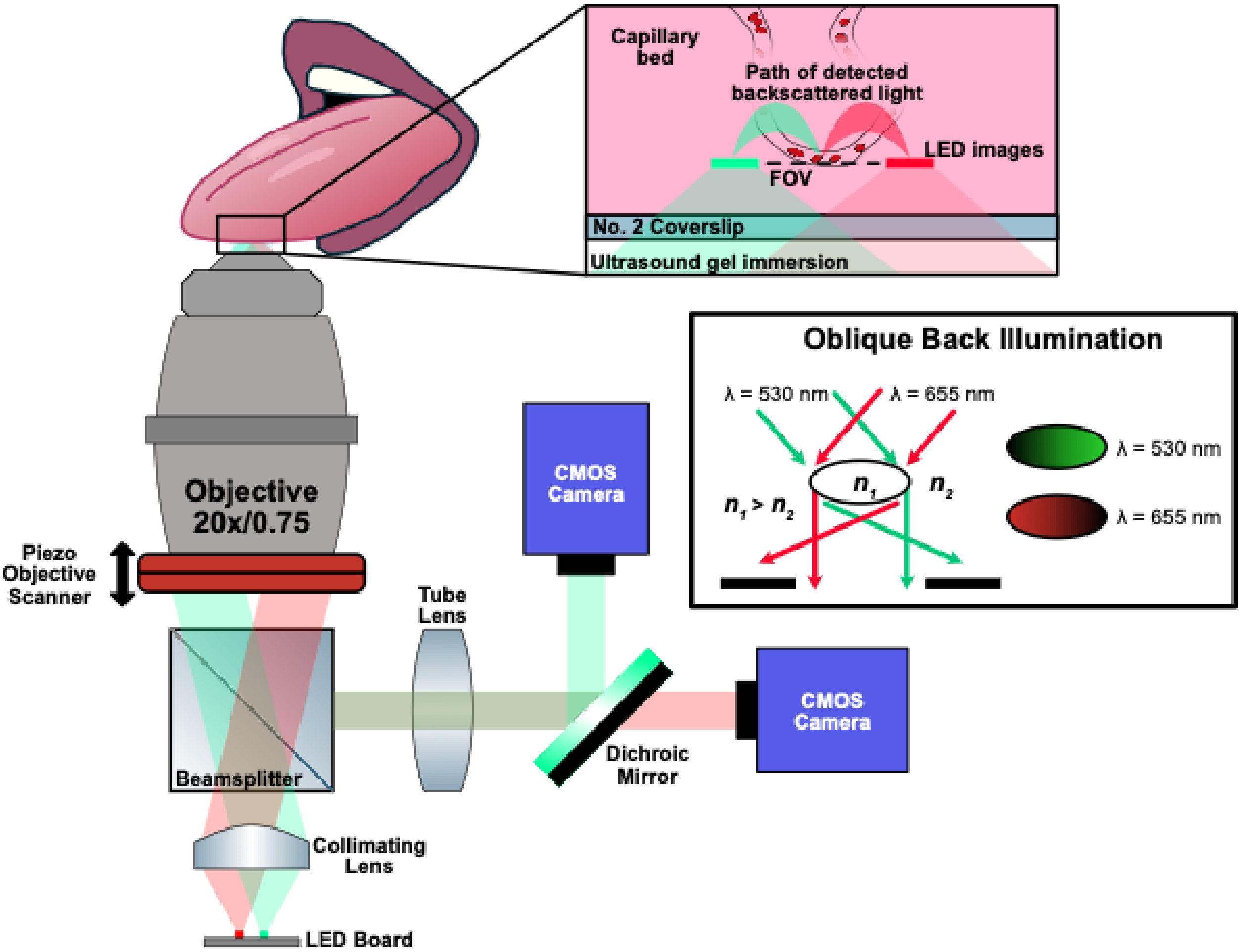


**Supplemental Figure 1. Optical system diagram.** Red (655nm) and green (530nm) LEDs are imaged onto the focal plane of the objective lens through a collimating lens and beamsplitter. The depth of the focal plane of the objective is adjusted with a piezo objective scanner. Light from the LEDs backscatters through the tissue and passes through the FOV at an oblique angle relative to the optical axis, producing contrast to lateral gradients in refractive index (phase). This light is reflected by the beamsplitter and refracted by a tube lens, forming an image on CMOS sensors.

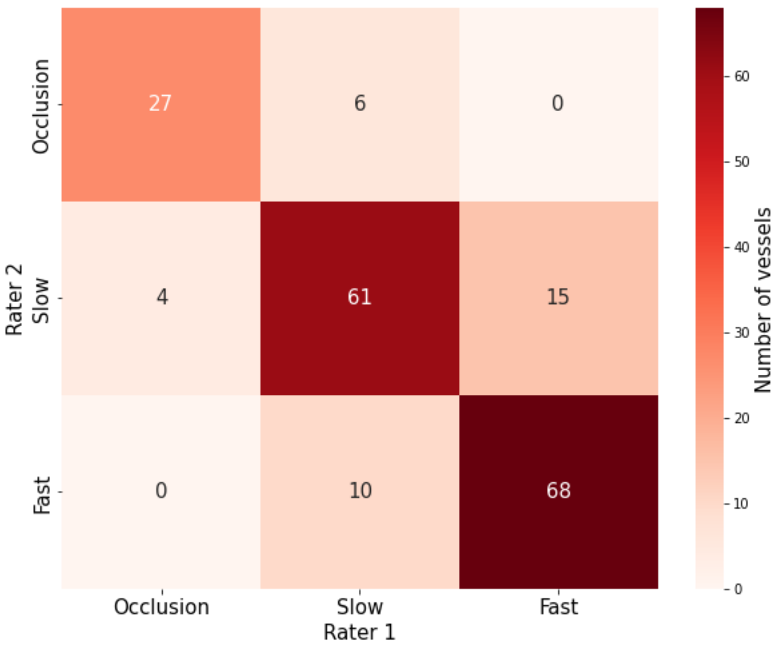


**Supplemental Figure 2. Confusion matrix of interrater agreement.** Comparison of flow categories of the same individual vessels assigned by two raters. 191 total vessels were observed in one randomly chosen subject with SCD from pre- and post-transfusion sessions combined.

**Video 1.** Fast flow example of vessel depicted in Figure 2a. Frames are stabilized and cropped. The green channel is shown on the left and the red channel on the right. Images are background subtracted and contrast enhanced using histogram-based equalization. Videos are 5x slowed down for visualization: frames were collected at 100 fps and played back at 20 fps.

**Video 2.** Slow flow example of vessel depicted in Figure 2b. Frames are stabilized and cropped. The green channel is shown on the left and the red channel on the right. Images are background corrected and contrast enhanced using histogram-based equalization. Videos are 5x slowed down for visualization: frames were collected at 100 fps and played back at 20 fps.

**Video 3.** Example of fast blood flow around an adhered cell depicted in Figure 3a. Frames are stabilized and cropped. The green channel is shown on the left and the red channel on the right.  Images are background corrected and contrast enhanced using histogram-based equalization. Videos are 5x slowed down for visualization: frames were collected at 100 fps and played back at 20 fps.

**Video 4.** Example of slow blood flow around adhered cell depicted in Figure 3b. Frames are stabilized and cropped. The green channel is shown on the left and the red channel on the right. Images are background corrected and contrast enhanced using histogram-based equalization. Videos are 5x slowed down for visualization: frames were collected at 100 fps and played back at 20 fps.

**Video 5.** Example of occlusion formation in the vessel depicted in Figure 3c. Frames are stabilized and cropped. The green channel is shown on the left and the red channel on the right. Images are background corrected and contrast enhanced using histogram-based equalization. Videos are 5x slowed down for visualization: frames were collected at 100 fps and played back at 20 fps.

**Video 6.** Example of an additional occlusion formation not shown in main Figures. Frames are stabilized and cropped. The green channel is shown on the left and the red channel on the right. Images are background corrected and contrast enhanced using histogram-based equalization. Videos are 5x slowed down for visualization: frames were collected at 100 fps and played back at 20 fps.
